# Potential magnitude of COVID-19-induced healthcare resource depletion in Ontario, Canada

**DOI:** 10.1101/2020.04.19.20071712

**Authors:** Kali Barrett, Yasin A. Khan, Stephen Mac, Raphael Ximenes, David MJ Naimark, Beate Sander

**Affiliations:** Institute of Health Policy, Management and Evaluation, University of Toronto, Toronto, Canada; University Health Network, Toronto, Canada; Toronto Health Economics and Technology Assessment (THETA) collaborative, University Health Network, Toronto, Canada; Sunnybrook Hospital, Toronto, Canada

## Abstract

**Background:** The global spread of coronavirus disease 2019 (COVID-19) continues in several jurisdictions, causing significant strain to healthcare systems. The purpose of our study is to predict the impact of the COVID-19 pandemic on patient outcomes and the healthcare system in Ontario, Canada.

**Methods:** We developed an individual-level simulation to model the flow of COVID-19 patients through the Ontario healthcare system. We simulated different combined scenarios of epidemic trajectory and healthcare capacity. Outcomes include numbers of patients needing admission to the ward, Intensive Care Unit (ICU), and requiring ventilation; days to resource depletion; and numbers of patients awaiting resources and deaths associated with limited access to resources.

**Findings:** We demonstrate that with effective early public health measures system resources need not be depleted. For scenarios considering late or ineffective implementation of physical distancing, health system resources would be depleted within 14-26 days. Resource depletion was also avoided or delayed with aggressive measures to rapidly increase ICU, ventilator, and acute care hospital capacity.

**Interpretation:** We found that without aggressive physical distancing measures the Ontario healthcare system would have been inadequately equipped to manage the expected number of patients with COVID-19, despite the rapid capacity increase. This overall lack of resources would have led to an increase in mortality. By slowing the spread of the disease via ongoing public health measures and having increased healthcare capacity, Ontario may have avoided catastrophic stresses to its health care system.

## INTRODUCTION

In December 2019, several patients in Wuhan, Hubei, China developed pneumonia, soon discovered to be due to a previously-unidentified coronavirus.(1,2) This virus, now termed severe acute respiratory syndrome coronavirus 2 (SARS-CoV-2) and the resulting clinical syndrome, coronavirus disease 2019 (COVID-19), has spread globally and was declared a pandemic on March 11, 2020.(3,4) As of April 12, there are more than 1.6 million confirmed patients with COVID-19, and more than 100,000 deaths reported worldwide.(5,6) The earliest rate of new infections was alarming, with the World Health Organization (WHO) reporting that the first 100,000 cases developed over three months, and the subsequent 100,000 cases developed over only 12 days.(7) In order to decrease spread of the disease, the global community has mobilized efforts to minimize social interactions.(8)

These interventions are imperative as the impact of COVID-19 is straining healthcare systems, especially critical care resources, worldwide.(9) Reports from China suggest that up to 20% of patients with COVID-19 require hospitalization,(10,11) with more than 26% of these patients admitted to the Intensive care Unit (ICU),(12,13) and nearly 50% of those requiring mechanical ventilation.(13) The situation is even more dire in Italy, where 12% of all patients with COVID-19 require ICU admission, compared to 5% in China.(9,14) Without measures to decrease the rate of spread, patients’ needs for critical care will overwhelm available resources.(15,16)

Predicting COVID-19 population spread and assessing interventions aiming to mitigate transmission are vitally important to ensure that healthcare systems are adequately prepared for this ongoing epidemic. Several epidemic models suggested that without an aggressive suppression strategy, i.e., simultaneous physical distancing, home isolation, quarantine, and university and school closure, the need for hospitalization and critical care would outstrip available resources. (17,18) While such transmission models are helpful to describe the magnitude of COVID-19 spread under different scenarios and provide high level estimates of impact on resource utilization, they do not adequately account for interactions between patients and the healthcare system.

Experience from China suggests that lower healthcare capacity is associated with a higher mortality rate.(19) Understanding how COVID-19 will affect healthcare resources is especially relevant in Canada, where there are 2.5 beds per 1,000 population; compared to 4.34, 3.84, 3.18 and 2.54 beds per 1,000 population in China, Australia, Italy, and the United Kingdom, respectively, as of 2018.(20) Fortunately, current data suggest that the rate of spread in Canada is lower than that seen in some of these countries, and this may be due to the early robust physical distancing policies enacted.

The goal of our study is to predict impact of COVID-19 on the acute care system and health outcomes in Ontario, Canada, as an example of a developed region with a mid-sized economy and a publicly-funded healthcare system, for a range of scenarios characterized by COVID-19 case predictions and acute care system capacity.

## METHODS

We developed a discrete-time, individual-level, health state transition model to forecast hospital resource utilization for symptomatic COVID-19 infected adults presenting to the hospital from the Ontario healthcare system perspective. The model incorporates a dynamic population, i.e., patients presenting daily to the hospital, and resource constraints: hospital ward beds, ICU beds, and ventilators. The primary outcomes are: 1) number of patients needing admission to the ward or ICU – with or without mechanical ventilation; 2) days to resource depletion of any of the resources: hospital ward beds, ICU beds, ventilators; 3) number of patients waiting for any of the resources per day and; 4) number of COVID-19-related deaths, stratified by whether or not patients received the needed resource. Outcomes were accrued over a time horizon of 60 days (2 months) and the cycle length (time steps) of the model was denominated in days.

## Model Structure

We simulated a dynamic population of adults (18 years and older) with symptomatic COVID-19 infection who arrive at the hospital emergency department (ED), where they will be sent home to self-isolate or be hospitalized (Figure 1). Individuals are admitted to a general medical ward or directly to ICU depending on disease severity, and some require invasive mechanical ventilation. Throughout the model, patients can either remain in their health state, recover, or die.

If any of the resources (ward beds, ICU beds, and ventilators) are unavailable, the patient is assumed to remain in their current state waiting for the resource to become available. For example, if a patient needs an ICU bed, and none are available, the patient will remain in the ED with no access to critical care resources until an ICU bed is available. Ventilators and ICU beds are freed up upon recovery or death of patients. Ward beds are freed up upon recovery of patients. Priority setting for ward bed resources is determined by the patient’s current location (i.e., ICU patients are prioritized over ED patients for ward beds) and, for other resources, priority is given to patients who have been waiting the longest since admission. All modelling and analyses were conducted using TreeAge Pro 2020 (TreeAge Software, Inc., Williamstown, MA).

**Figure 1.**
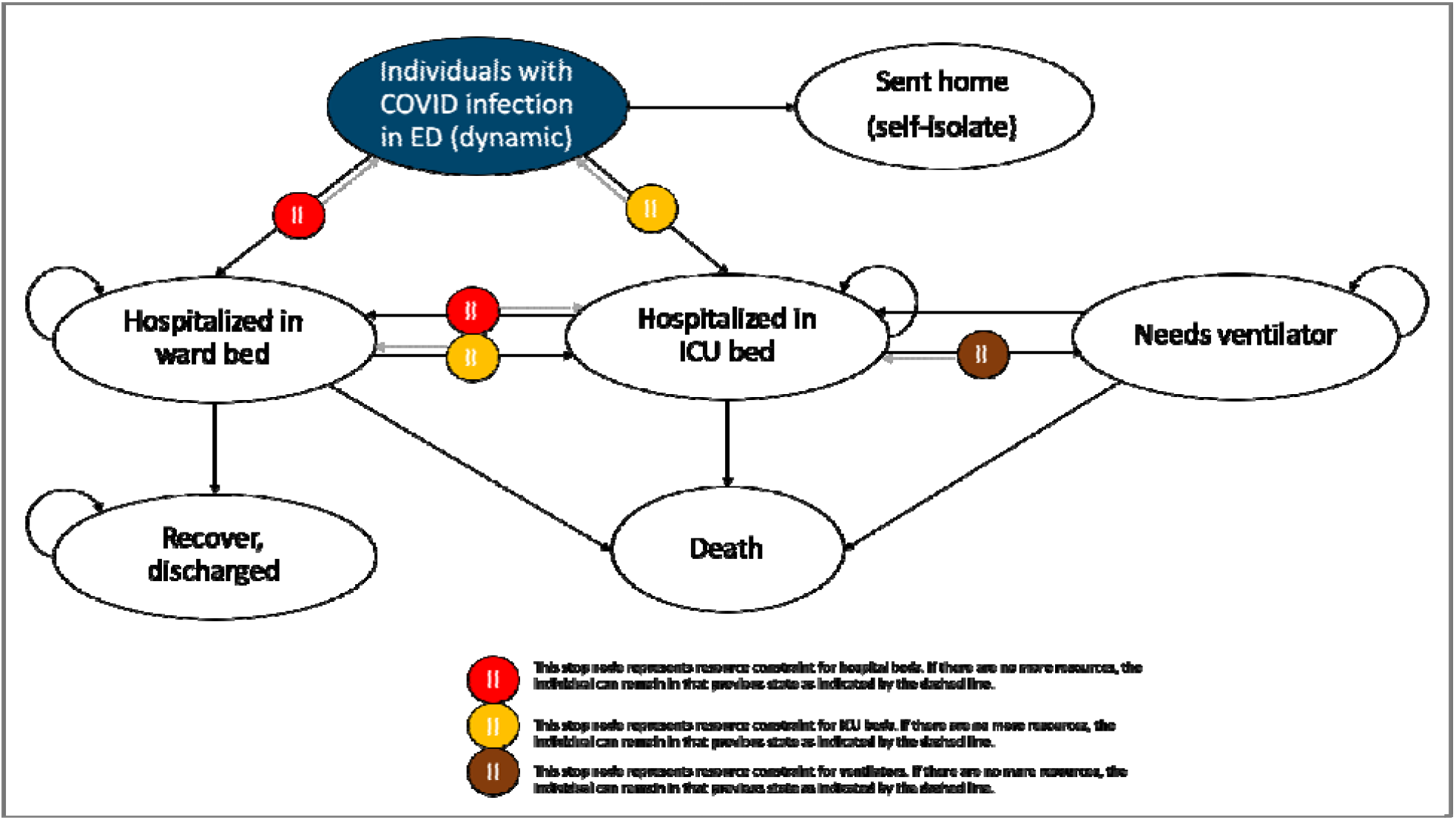
Model Schematic.

## Key Parameters and Data Sources

### COVID-19 disease history

Data for the model was extracted from the evolving literature on COVID-19, supplemented by expert guidance (Table 1).

**Table 1.** Key Variables

| **Variables** | **Base-case Value** | **Sources** |
| --- | --- | --- |
| Number of infected subjects, per day, by strategy | See Fig. 2 | Estimated as described in text |
| Probability needing hospitalization | 0.18 | Public Health Agency of Canada estimate (21) |
| Probability of needing ICU level care given hospitalization | 0.48 | Calibrated based on Public Health Agency of Canada estimate (21) |
| Probability of ICU patients needing ventilation | 0.78 | Critical Care Services Ontario (CCSO) estimate on April 13, 2020 (22) |
| Probability that patients on the ward deteriorate and need ICU | 0 | Assumption |
| Length of stay, ward (no ICU admission/prior to ICU admission) | 17 days | Bellani 2016 (24) |
| Length of stay, ICU (with/without ventilation) | 11 days | Bellani 2016 (24) |
| Length of stay, ward post-ICU | 6 days | Bellani 2016 (24) |
| Probability of death ward patients | 0 | Wu 2020 (23) |
| Probability of death, ICU-patients | 0.35 | Bellani 2016 (24) |
| Probability of death, ventilated patients | 0.35 | Bellani 2016 (24) |
| Probability of death, patients waiting for vent | 1.0 | Assumption |

We assumed that the probability of hospitalization in Ontario was similar to crude national hospitalization estimates of 18%.(21) In the absence of good data, ICU admission given hospitalization for Ontario was estimated to be 48% by calibrating to observed ICU admissions. The Ontario estimate for ICU admissions requiring ventilation was 78%, as reported by Critical Care Services Ontario on April 13, 2020.(22) In keeping with the current literature, we assumed that only patients requiring ICU admission have a risk of death.(23) We assumed the same mortality risk for patients who require non-ventilated intensive care but cannot access ICU beds due to resource constraints. We assumed that patients who need mechanical ventilation but cannot access it will die within that day. The probability of death in the ICU (for individuals requiring, and not requiring mechanical ventilation) was based on the proportion of deaths from Bellani and colleagues of 35%,(24) over the mean length of ICU stay of 11 days for patients (and a mean hospital length of stay of 17 days) with moderate acute respiratory distress syndrome (ARDS). We used length of stay and mortality estimates of moderate ARDS patients as they are considered clinically similar to COVID-19 cases in the ICU per expert guidance.

### The effect of COVID-19 on healthcare resource availability in Ontario

In the absence of effective treatments for COVID-19 per se, desirable health outcomes, on a population level, can be maximized by reducing the number of cases through public health measures (e.g., physical distancing, testing, and isolation) and by increasing system capacity (freeing up existing resources or adding new additional resources). We therefore explore a range of scenarios considering COVID-19 spread and system capacity. We present forecasts for 9 scenarios: three possible epidemic trajectories (labelled ‘A’, ‘B’ and ‘C’) and three resource availability scenarios (base case [BC], 1, and 2) described below.

### Epidemic trajectories

We modeled three potential scenarios of the possible epidemic trajectory in Ontario starting on March 6, 2020 when Ontario had reported over 100 cumulative COVID-19 cases (actual: 109 cases).

A. <u>“South Korea scenario”</u>: started at 104 cumulative cases, aligned to Ontario’s starting date. We used observed daily incidence for South Korea for 50 days (reported data up until April 13^th^, 2020 (5)).
B. <u>“Expected scenario”</u>, started at 109 cumulative cases on March 6, 2020. We used observed data from Ontario’s integrated Public Health Information System (iPHIS) up until March 30 to account for reporting delays, applied the mean daily increase in cumulative cases per day observed in the seven days prior (7.1%), and assumed a peak at April 7, then applied a 5% daily decrease (contraction rate) in new cases per day (incidence) until day 60. The contraction after April 7 represents the effect of public health measures enacted in Ontario in mid-March.
C. <u>“Italy Scenario”</u>, started at 155 cumulative cases which was best aligned to Ontario’s starting date of March 6, 2020. We used observed daily incidence for Italy for 50 days (reported data up until April 13^th^, 2020).(5)

Daily numbers of expected cases for each scenario are shown in Figure 2.

**Figure 2:**
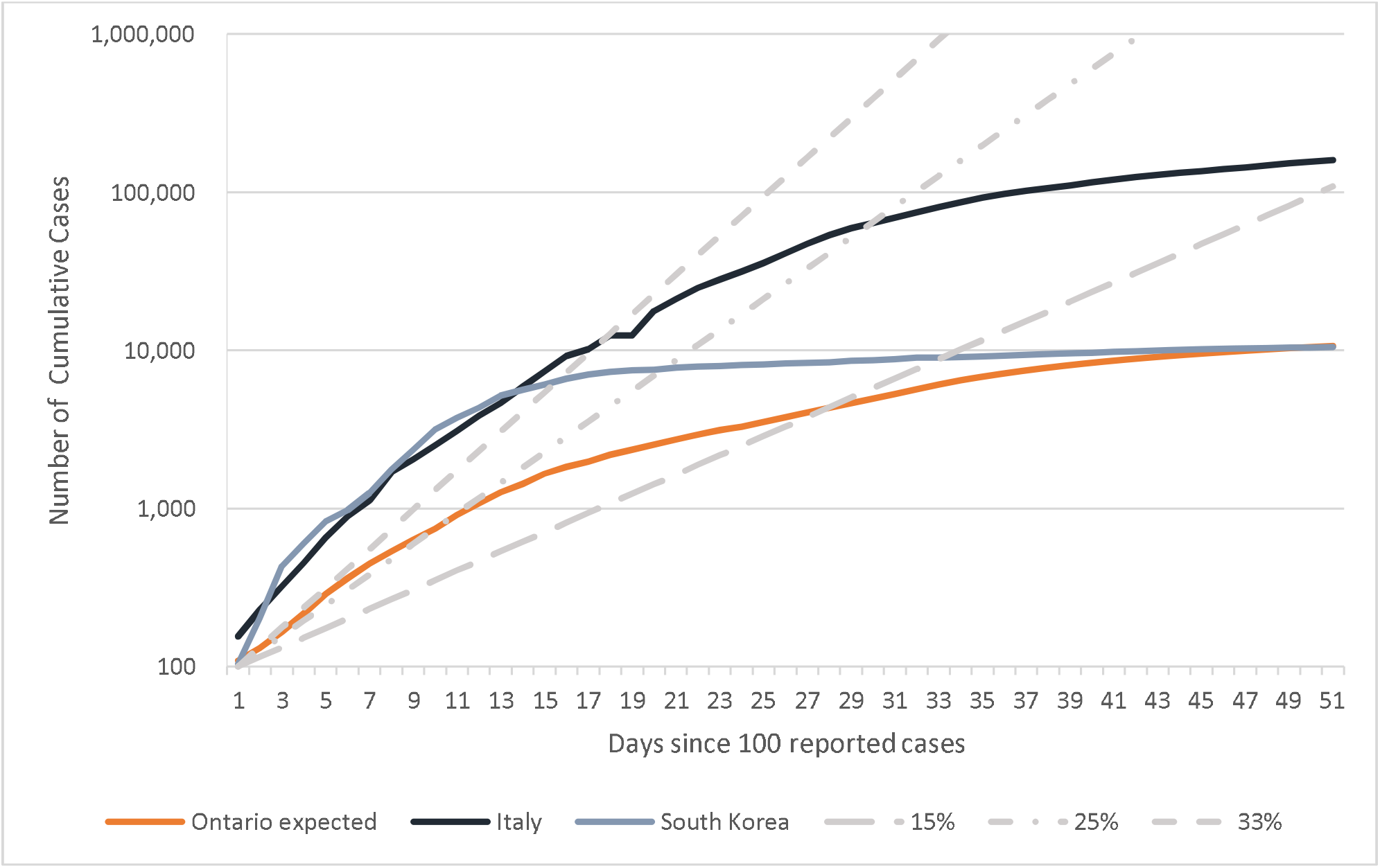
Predicted Cumulative Number of New Cases over Time.

### Hospital resource scenarios

We consider ward beds and ICU beds with or without a ventilator. For a patient to receive mechanical ventilation, both a ventilator and ICU bed need to be available. We assume all available resources are appropriately staffed.

For our base case (BC), the number of available ventilator beds at the start of the simulation represents the number of existing ventilators in Ontario. We assumed that approximately 25% of total existing Ontario ventilator beds (328 out of 1,311), and non-ventilator ICU beds (186 out of 742) would be available to accommodate COVID-19 patients based on expert judgement. We assumed a hospital ward bed availability rate of 20% for COVID-19 patients, based on expert judgement (4,000 out of 20,000 acute care beds). In the base-case, we assumed that no additional beds or ventilators would be made available for COVID-19 patients. Using this base case, we simulated the three epidemic trajectories: “South Korea scenario” (BC-A), “Expected scenario” (BC-B), and “Italy scenario” (BC-C).

We conducted two expanded resource availability scenarios to explore the impact of potential resource increases on time to resource depletion, patients waiting for each resource, and mortality. We explored potentially achievable as well as an extreme scenarios:

1. Employment of surge capacity: The proportion of beds available to COVID-19 patients is 25% of the total existing ICU beds (ventilated and non-ventilated), and 20% of ward beds (as in the base case scenario), plus additional surge capacity of 502 ventilator beds, 351 non-ventilator ICU beds, and 1,351 ward beds (total 830 ventilator beds, 537 non-ventilator ICU beds, and 5,351 ward beds). This scenario explores the effect of increasing capacity by maximizing currently available resources through a reduction in clinical activity such as elective surgeries and activating existing surge capacity protocols (personal communication Ontario Health). We refer to these scenarios as 1A, 1B, and 1C.
2. A vast increase in health system capacity through unconventional hospital space: 2,000 ventilator ICU beds (∼ 50% more than existing capacity), 1,000 non-ventilator ICU beds (∼ 50% more than existing capacity), and 10,000 ward beds (50% of existing capacity) would be available to COVID-19 patients. We refer to these scenarios as 2A, 2B, and 2C.

A summary of all analyses is presented in Table 2.

**Table 2.** Summary of base-case and scenario analyses

|  |  | Predicted Number of Cases |  |  |
| --- | --- | --- | --- | --- |
|  |  | A<br>“South Korea<br>Scenario” | B<br>“Expected Scenario” | C<br>“Italy Scenario” |
| Capacity | **Base Case (BC)**<br>• 328 ventilator ICU beds<br>• 186 non-ventilator ICU beds<br>• 4,000 ward beds | BC-A | BC-B | BC-C |
|  | **Expanded 1**<br>• 830 ventilator ICU beds<br>• 537 non-ventilator ICU beds<br>• 5,351 ward beds | 1A | 1B | 1C |
|  | **Expanded 2**<br>• 2,000 ventilator ICU beds<br>• 1,000 non-ventilator ICU beds<br>• 10,000 ward beds | 2A | 2B | 2C |

### RESULTS

## Base-case analysis

In Table 3, we summarize all findings for the base-case in all three epidemic trajectories.

**Table 3.** Summary of resource constraints and mortality

|  | Days Until Resource Unavailable |  |  | Deaths |  |
| --- | --- | --- | --- | --- | --- |
|  | ICU Beds | Ventilators | Ward Beds | While waiting for needed resources | Those receiving needed resources |
| **Base-case** |  |  |  |  |  |
| A (S Korea) | Never | Never | Never | 0 | 321 |
| B (Expected) | Never | Never | Never | 0 | 392 |
| C (Italy) | 17 | 14 | 26 | 11,174 | 945 |
| **Scenario 1** |  |  |  |  |  |
| A (S Korea) | Never | Never | Never | 0 | 321 |
| B (Expected) | Never | Never | Never | 0 | 392 |
| C (Italy) | 25 | 19 | 28 | 7,413 | 2,314 |
| **Scenario 2** |  |  |  |  |  |
| A (S Korea) | Never | Never | Never | 0 | 321 |
| B (Expected) | Never | Never | Never | 0 | 392 |
| C (Italy) | 34 | 27 | Never | 2,049 | 4,350 |

### Scenario BC-A

Our base-case simulation of the epidemic scenario A (“South Korea”) predicted a total of 10,433 COVID-19 cases, of which 2,295 required hospital admission over 60 days. Following this epidemic trajectory, we predict that Ontario’s ICU beds, and ventilator resources would be used extensively between days 10 and 20 due to the rapid increase in COVID-19 cases, but never completely depleted. Ward beds would also not be depleted (Figure 3). In this scenario, 321 patients died from COVID-19 despite receiving appropriate care and hospital resources.

**Figure 3.**
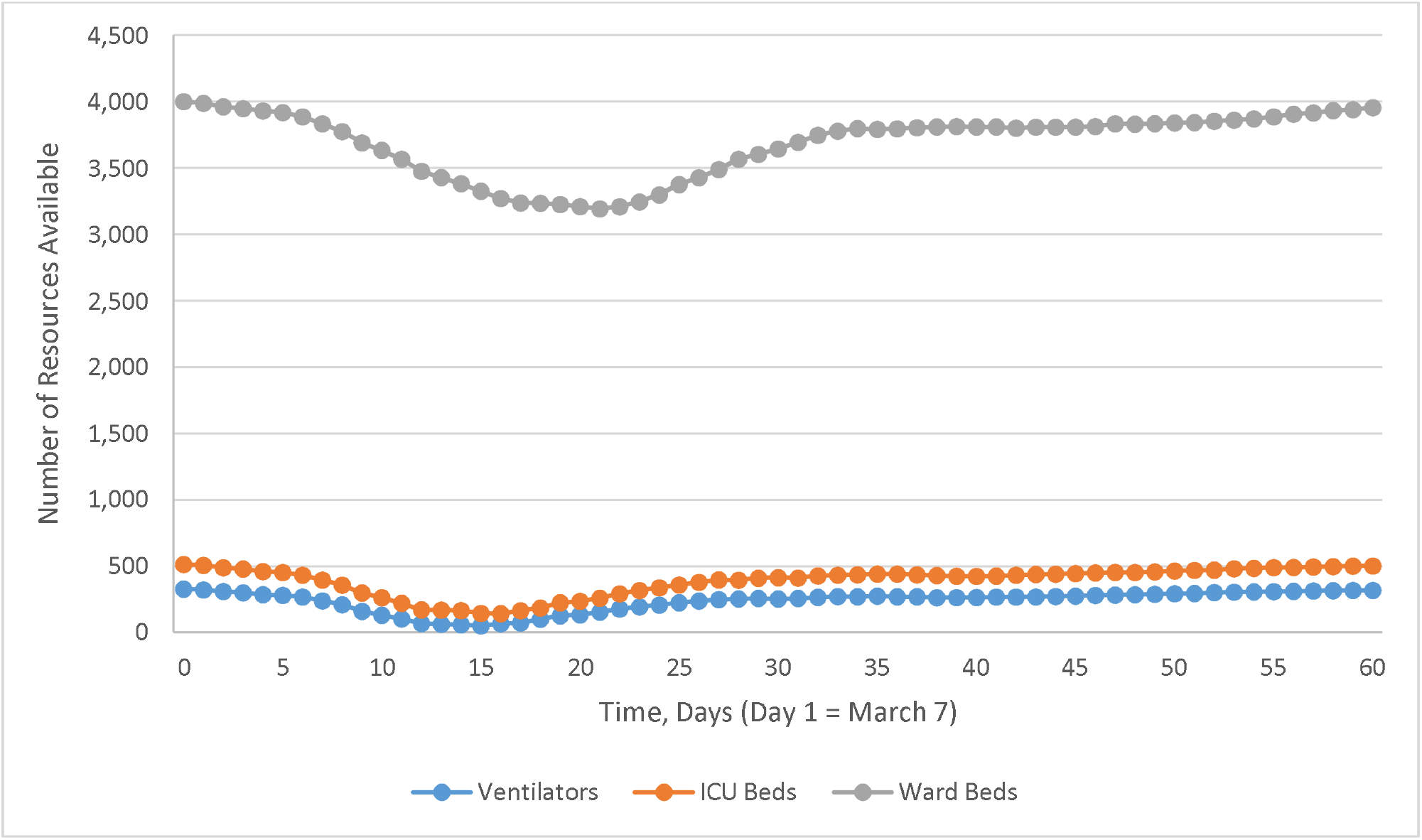
Resource depletion for Scenario BC-A (Base case resources, South Korea trajectory)

### Scenario BC-B

In the base-case analysis for epidemic scenario B (“Expected”), our simulation predicted a total of 11,705 COVID-19 cases, of which 2,575 required hospital admission over 60 days. Using the observed data for Ontario, where physical distancing measures were implemented on March 15, and projecting an April 7 peak for incidence of COVID-19 cases, Ontario’s ICU bed and ventilator resources would not be depleted (Figure 4). In this scenario, 392 patients died from COVID-19 despite receiving appropriate care and hospital resources.

**Figure 4.**
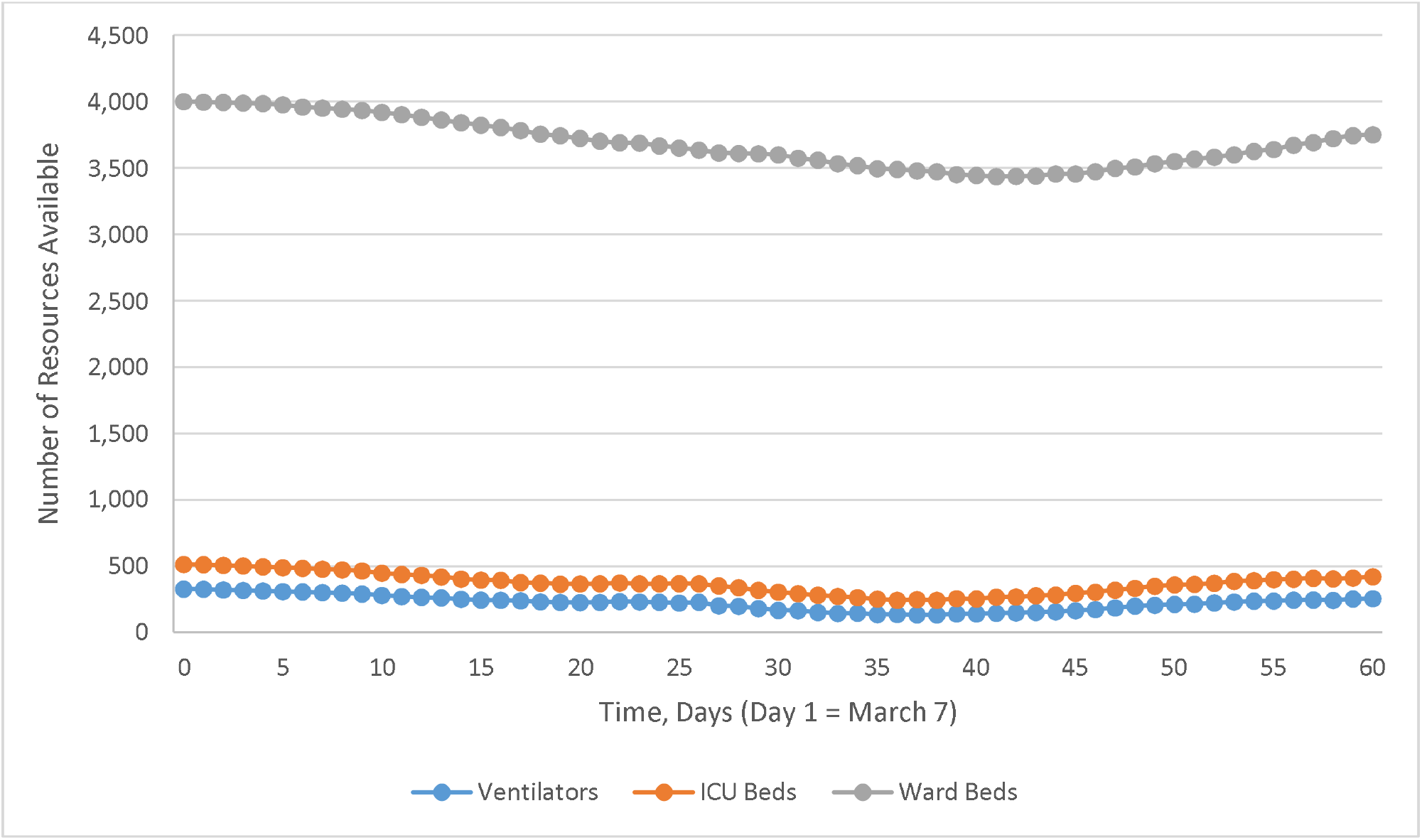
Resource depletion for Scenario BC-B (Base case resources, expected trajectory)

After calibration, the projected ICU and ward bed occupancy using this scenario compared favourably to the observed data in Ontario (Figure 5).

**Figure 5.**
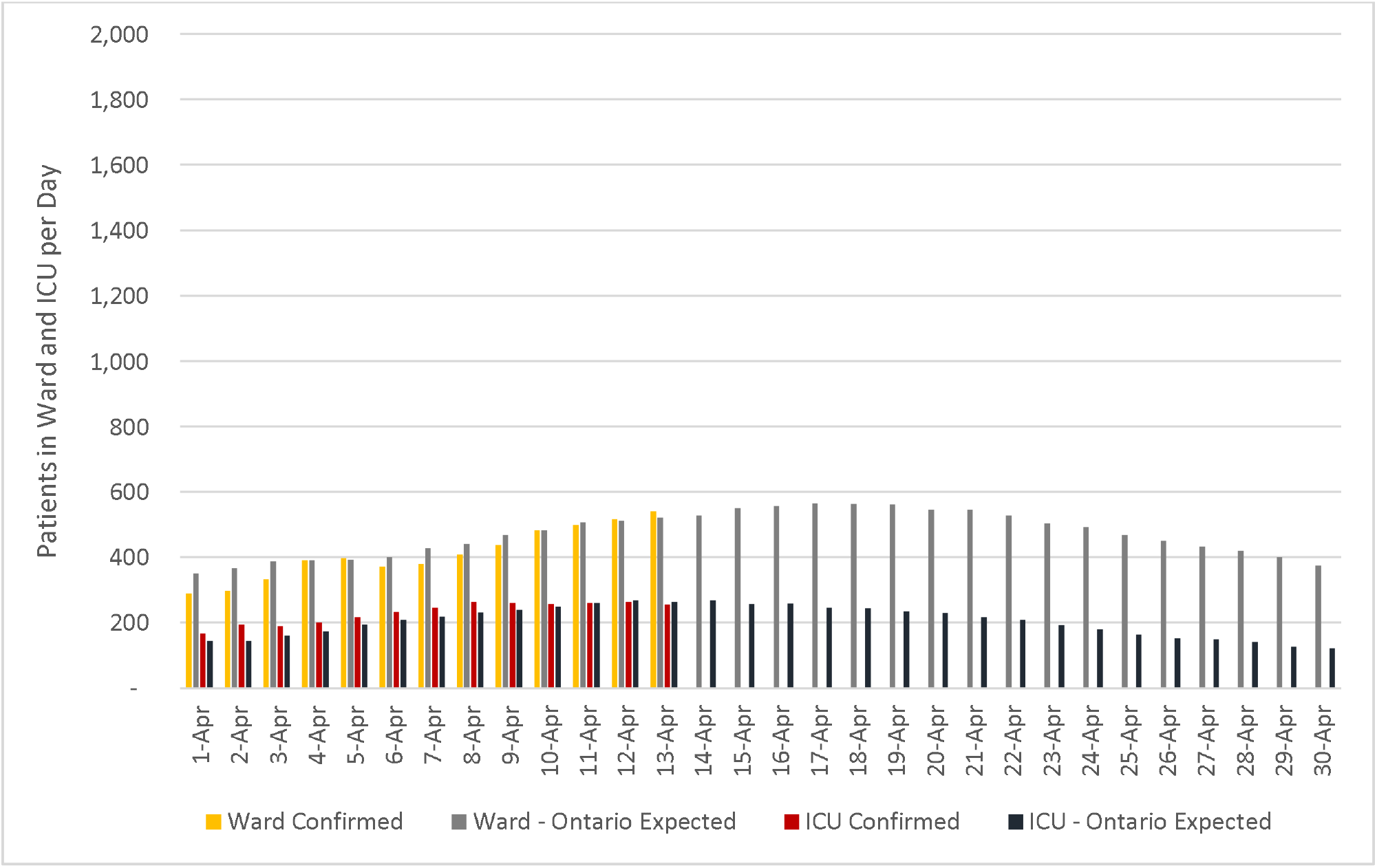
Ward and ICU occupancy for COVID-19 patients over time in Ontario.

### Scenario BC-C

In the base-case analysis for epidemic scenario C (“Italy”), our analysis predicted a total of 159,361 COVID-19 cases, of which 35,059 required hospital admission over 60 days. Following this epidemic trajectory, Ontario’s ICU bed and ventilator resources would have been depleted in approximately 17 and 14 days, respectively, from the start. Ward beds would have been full and unable to accommodate new patients in approximately 4 weeks (26 days) (Figure 6). In this scenario, where there are more daily cases of COVID-19-infected patients, and an earlier date of resource depletion, we observed the greatest number of patients dying while waiting for appropriate care, 11,174, compared to 945 patients dying from COVID-19 who had received appropriate care.

**Figure 6.**
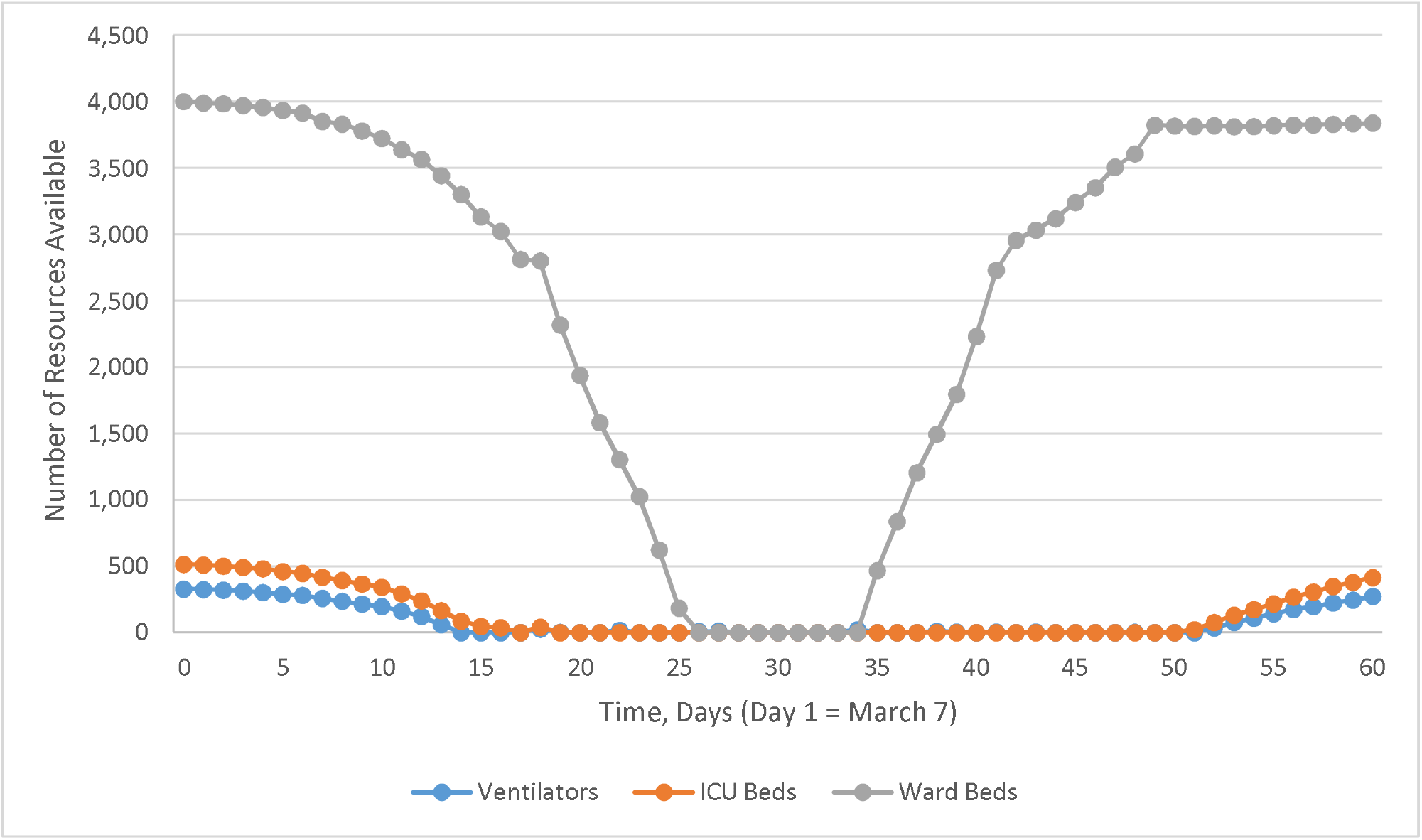
Resource depletion for Scenario BC-C (Base case resources, Italy trajectory)

## Resource expansion scenarios

### Days Until Resources are Depleted

Expanded capacity scenarios 1 and 2 were simulated for each epidemic trajectory scenario A, B and C (Table 3). For epidemic scenario A and B, the change in ICU bed and ventilator capacity did not materially affect the results in terms depletion for the ward beds, ICU beds and ventilators compared to the base case resource scenario: these resources were never completely depleted.

In contrast, for epidemic scenario C, simulation of capacity scenarios 1 and 2 (Figures 7 and 8, respectively) showed that increased ICU beds and ventilator resources delayed but did not eliminate complete ICU bed and ventilator depletion. For example, in Scenario 1C where there is an increase in the number of ventilators, ICU beds and ventilators are depleted at approximately 25 and 19 days, respectively, instead of approximately 17 and 14 days from the start date.

**Figure 7.**
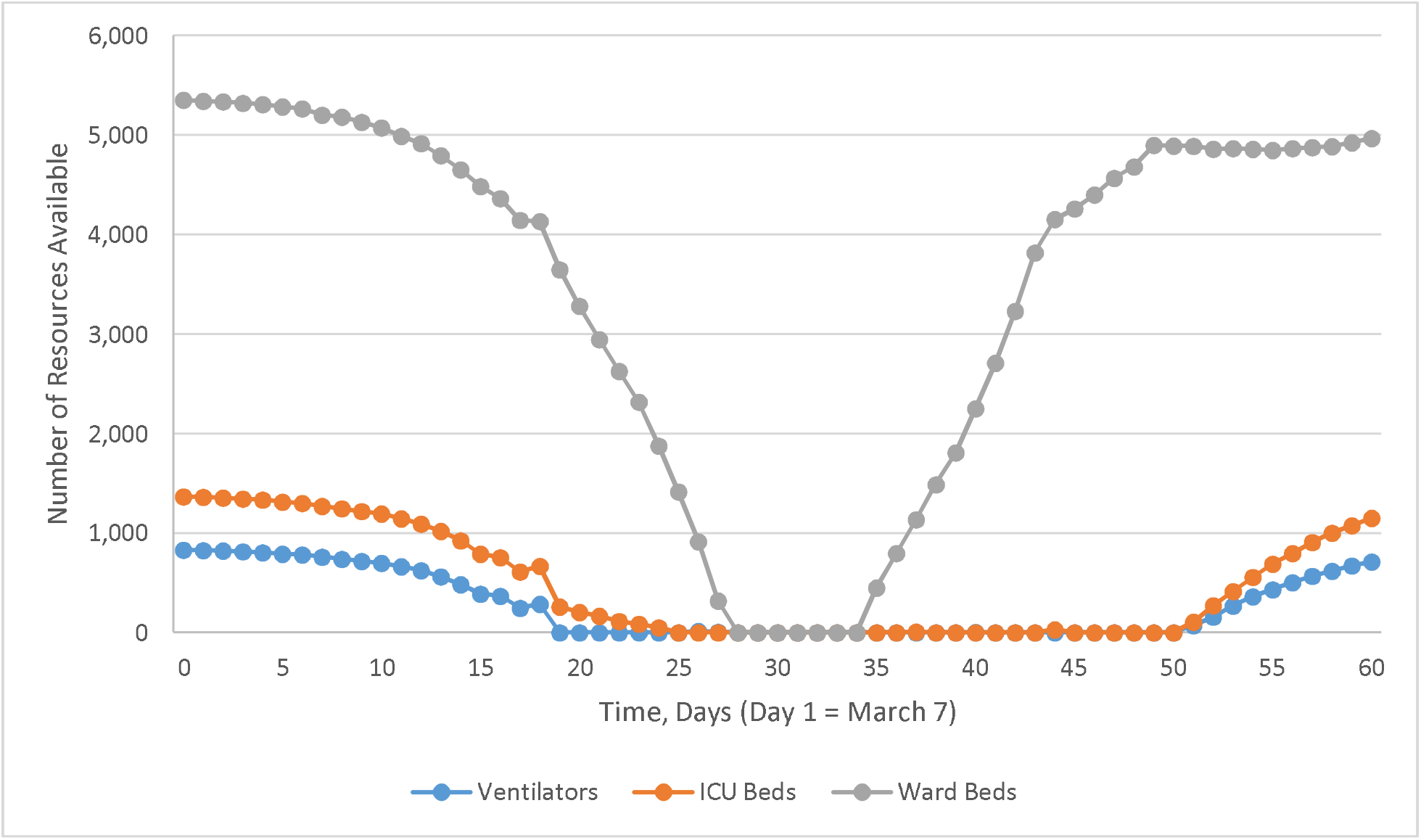
Resource depletion for Scenario 1-C (Surge capacity expansion, Italy trajectory)

**Figure 8.**
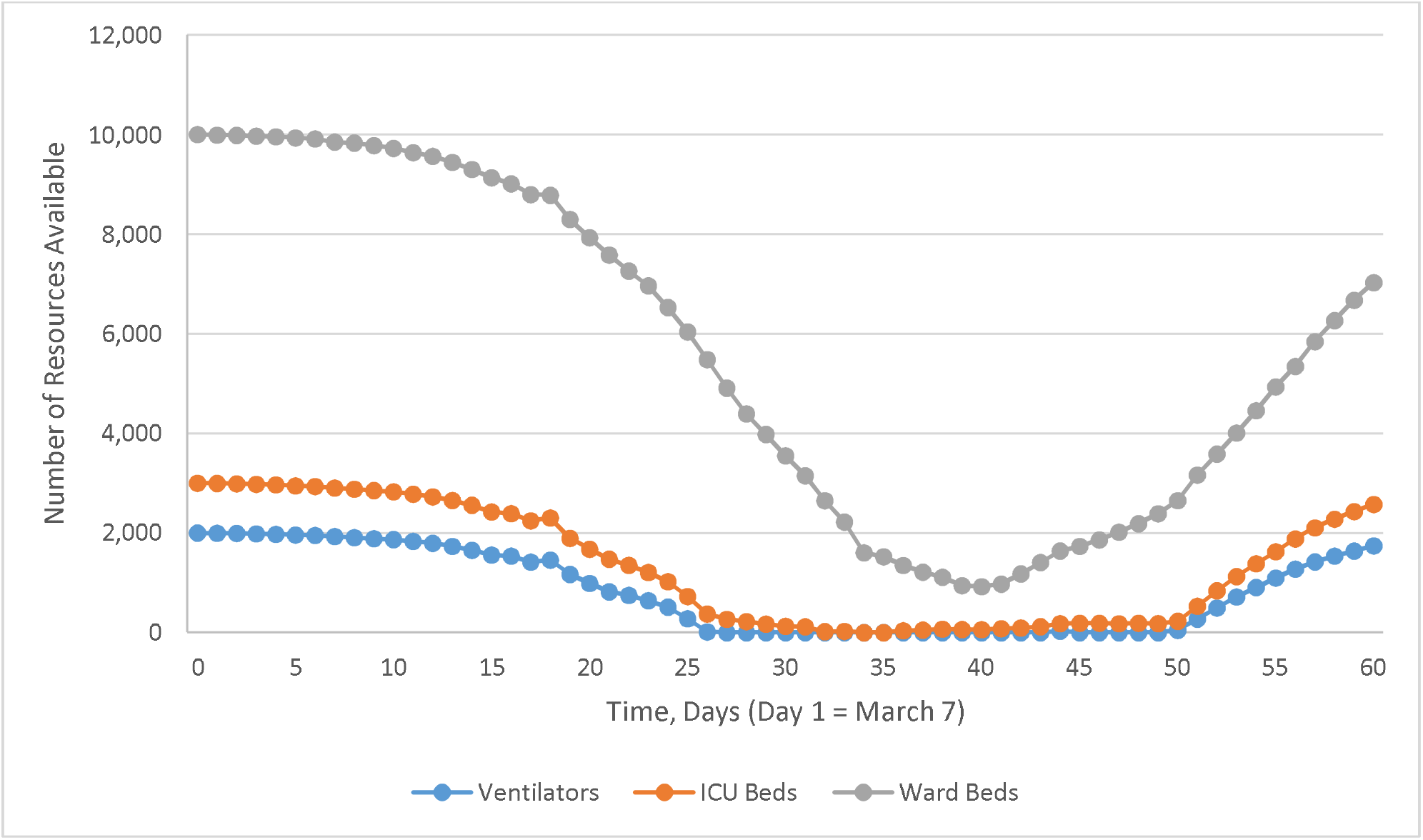
Resource depletion for Scenario 2-C (Massive capacity expansion, Italy trajectory)

### Mortality

Deaths in patients waiting for resources were substantially higher in the base-case scenario C (BC-C) than in base-case scenario B (BC-B) due to increased numbers of COVID-19-infected patients. However, as the number of resources increased in Scenario 1 and 2, the number of deaths in patients waiting for resources decreased for scenario C, while the number of deaths for those receiving needed resource increased (Table 3).

## DISCUSSION

Since December 2019, worldwide number of cases of COVID-19 has continued to rise and the daily rate of new cases is still increasing in several jurisdictions.(6) The trends in Canada and Ontario mirror what has been reported in the rest of the world.(25,26) Reports from other jurisdictions with COVID-19 outbreaks suggest that healthcare resources are indeed in short supply (27,28) and that planning for rationing of ICU resources is underway.(29)

As the earlier transmission models suggest, the burden on the healthcare system can be decreased by public health measures that aim to slow the spread and delay the epidemic peak of COVID-19 infections. Furthermore, aggressive isolation and quarantine approaches have already been effective in decreasing new infections (30) and preventing outbreaks.(31)

Our results demonstrate that physical distancing and other public health measures are effective at reducing health system burden. They also demonstrate that public health measures used in conjunction with efforts to rapidly increase health system capacity for acutely ill patients significantly reduces or delays the likelihood of health system collapse and resource depletion. We calibrated our predictions using observed ICU occupancy, which has less uncertainty than reported case incidence. The latter is affected by reporting delays and changing testing policies. As of April 13, 2020, predictions regarding ICU occupancy from the expected epidemic trajectory scenario aligns well with current observed data in Ontario. These data demonstrate stable ICU occupancy rate for confirmed COVID-19 patients over the last week and suggest that public health interventions introduced in March in Ontario are starting to have a measurable effect. In contrast, had Ontario not taken these steps and, instead, followed an epidemic trajectory resembling Italy’s, even with massive expansion of resource capacity, critical care in the Province would have collapsed quickly and catastrophically.

Furthermore, our results show that deaths while waiting for resources can be avoided through implementation of public health measures. In contrast, without such measures, resource expansion, even on a massive scale, would not have been sufficient to prevent substantial numbers of these deaths. Our results therefore strongly support continued aggressive public health measures aimed at slowing the spread of the infection in order to decrease the epidemic size and minimize deaths.

Our study has several limitations. The model currently relies on forecasting COVID-19 cases based on reported data from Ontario and other countries, projections, and assumption on physical distancing effectiveness. Also, since we assumed that patients requiring ward beds will not die from COVID-19 based on the current literature, we may underestimate the number of overall deaths. Given current data, we assumed a fixed number of ward beds, ICU beds, and ventilators. However, we simulated scenario analyses to demonstrate the impact of increasing availability of existing resources as well as adding additional resources. Currently available data that informs our acute care and ICU length of stay estimates, or the effect of insufficient resources on mortality is limited for COVID-19 patients. Priority setting is modeled so that ICU patients requiring ward beds have access to any ward bed ahead of incoming patients, regardless of their wait time. Other resources are available based on time of admission and not time since the resource was first needed. Our model does not incorporate COVID-19 transmission in the hospital, potentially underestimating resource need. Lastly, we do not explicitly consider health human resource constraints or the availability of adequate personal protective equipment, and assume that all hospital resources are appropriately staffed and have necessary supplies. Our study considers epidemic growth and resources for the whole of Ontario and does not consider regional differences within the province.

Our study has several strengths. In addition to modeling different disease transmission scenarios, we have also included a range of potential scenarios for an increase in healthcare capacity. To our knowledge, ours is the first study to explicitly quantify and forecast these effects. Our study is also the first to model patient flow through the healthcare system under these different scenarios in order to estimate resource utilization and availability at each level of care. These approaches have allowed us to predict the particular effect of resource depletion on rates of mortality. Another strength that lends to the validity of our study is the incorporation of observed increases in daily case count data from countries with both early and aggressive public health measures and those in which these efforts were lacking. The validity of our findings is bolstered by our use of published reports for clinical outcomes and resource utilization. Finally, we obtained current estimates of local healthcare resources, and used data from governmental and healthcare system proposals for increasing these resources in our estimates of potential capacity.

## CONCLUSIONS

Our model suggests that healthcare resources in Ontario currently appear to be adequate to manage the increasing number of patients with COVID-19. This result occurred because of public health measures that have aimed to reduce the rate of disease transmission in conjunction along with aggressive and rapid efforts to increase health system capacity. Results from the counterfactual scenario in which these steps had not been taken are sobering. They point to the need to maintain rigorous public health interventions in the near term.

## Data Availability

Data used in this manuscript can be referenced in the text.

